# Low prevalence of scabies and impetigo in Dakar/Senegal: a cluster-randomised, cross-sectional survey

**DOI:** 10.1101/2023.05.23.23290443

**Authors:** Andreas Hansmann, Genevia Wamba Lékémo, Chiaka Fomba, Jade Kaddoura, Ramatoullaye Toure, Assane Diop, Maodo Ndiaye, Olivier Chosidow, Michael Marks, Fatimata Ly

**Author notes:** Ampersand These authors are joint first authors (Andreas Hansmann, Genevia Wamba Lékémo). Asterisk These authors are joint senior authors (Olivier Chosidow, Michael Marks, Fatimata Ly). Hôpital Pitié-Salpêtrière, Paris, France (Olivier Chosidow).

## Abstract

**Background:** Scabies, a parasitic infection caused by *Sarcoptes scabiei var. hominis, is* a public health problem with significant morbidity worldwide, particularly in low-resource countries. Impetigo, a complication of scabies infection, is a risk factor for sepsis, glomerulonephritis and possibly acute rheumatic fever. Currently, the majority of epidemiological data has been collected in rural populations in the Pacific with limited applicability to urban populations in sub-Saharan Africa, where scabies is also believed to be a problem. To inform future public health programs more reliable information about the burden of disease is required.

**Methodology/Principal Findings:** In July/August 2022 we conducted a cross sectional, cluster-randomised, household survey in Pikine/Dakar using the ‘International Association for the Control of Scabies (IACS)’ criteria to diagnose scabies and impetigo. All participants underwent a standardised clinical examination by post-graduate dermatology students. For those diagnosed with scabies an age-adapted ‘Dermatology Life Quality Index (DLQI)’ questionnaire was filled.

We recruited and examined 1697 participants detecting 27 cases of scabies (prevalence: 1.6%, 95% CI 0.8-3.2), mostly in school aged children. 10 participants suffered from impetigo (prevalence: 0.6%, 95% CI 0.3-1.3), 5 of which were dually infected with scabies. Risk factors for scabies infection were young age, male gender and koranic school attendance. Scabies had a large effect on the lives of 7 out of 22 cases (31.8%), who had DLQI questionnaires filled.

**Conclusions/Significance:** This study adds to the mapping of the burden of scabies across Africa to support public health action. With a low prevalence of scabies that is concentrated amongst poor households and children attending koranic schools a focused public health approach targeting koranic schools and poor households seems to be most appropriate in this community.

**Author’s Summary:** Caused by a mite, scabies is a very common skin infection transmitted between humans. It most commonly affects children in tropical countries and causes a severe itch leading to emotional distress and sometimes bacterial infections than can lead to severe illness.

Despite efforts in the past to reduce the burden from scabies at a population level a lack of knowledge about the distribution of the parasite hampers the design and implementation of larger control programs. In Africa, little is known about the burden and distribution of scabies, particularly in urban centres.

This study surveyed inhabitants of Dakar for scabies and bacterial skin infections. We randomly selected groups of houses in a low socio-economic status neighbourhood and invited all inhabitants of these houses for an interview and for a brief skin examination.

Of 1697 participants only 27 (1.6%) were found to have scabies, 5 of whom also suffered from impetigo, a bacterial skin infection. Most people with scabies were pupils in koranic schools suggesting they may be the setting of ongoing transmission. Further studies need to find reasons for the high burden amongst these pupils and how best to reduce the burden in these schools.

## 1. Introduction

Scabies is a parasitic skin infection caused by the *Sarcoptes scabiei var. hominis* mite. It is estimated that worldwide more than 200 million people live with scabies and 455 million new cases occur every year accounting for 4.5 million DALYs. (1) (2) (3) Whilst scabies is endemic worldwide, the largest burden is recorded in low-income, tropical countries with children and the poorest in society disproportionally affected. (4) (5)

Scabies is transmitted predominantly through close skin-to-skin contact within households or in institutional settings. (6) Female mites burrow into the skin of the human host depositing eggs and excrement. (7) Intense itch occurs from three weeks after infection leading to distinctive skin eruptions. The intense itch, scratching and skin changes can lead to psychosocial consequences including shame, stigma, isolation, sleep disturbance and absenteeism from school or work. (8)

Scabies can also have serious health outcomes via bacterial superinfection (‘impetiginisation of scabies’) with *S. pyogenes* or *S. aureus*. This can contribute significantly to the burden of local and systemic bacterial infection, including necrotizing soft tissue infection, sepsis, post-streptococcal glomerulonephritis (9) (10) (11) (12) and possibly rheumatic fever and rheumatic heart disease (13) (14). Because scabies is so common and can cause potentially severe health outcomes through impetiginisation there has been renewed effort to control the disease.

In 2017 WHO added scabies to its list of Neglected Tropical Diseases (NTDs) and noted that mapping of the disease’s prevalence needs to occur before large-scale activities associated with scabies prevention and control can begin. (15) To identify communities who would benefit from this intervention WHO advocates for a rapid mapping of scabies prevalence in suspected high-burden countries. (16). To standardize efforts the International Alliance for the Control of Scabies (IACS) published consensus criteria for the diagnosis of scabies in 2018. (17) (18). Initial evaluations have demonstrated the validity of these criteria. (19) (20) (21).

Geographically, most population prevalence surveys have been conducted in the Pacific and have frequently recorded very high prevalence rates amongst the general populations. (22) (23) (24) (25). In Africa, a community-wide survey in rural Malawi found scabies and impetigo prevalence rates of 15% and 7% respectively. (26) In the Amhara Region of Ethiopia a population-based survey amongst children aged 5 to 14 years reported a scabies prevalence of 11%. (27). In peri-urban Monrovia a population-based survey of scabies and impetigo recorded a scabies-prevalence of 9%, half of which was severe (52%). 1% of the population suffered from impetigo. (21) Surveys in Guinea Bissau and the Gambia have found a prevalence of scabies of 5 and 15 % respectively amongst children. (28) (29)

In Senegal, to our knowledge, no large population-based surveys of scabies or impetigo has been conducted in the past, though anecdotal evidence suggests a high burden of disease. A recent survey of 15 Koranic schools, called ‘daaras’, in Dakar found a prevalence of 7.3% amongst pupils, with a range from 2 to 31% between individual institutions. (30) Daaras are residential schools where students learn the Koran, at times in very crowded circumstances. Surveys in prisons and dermatologists practicing in public hospitals in Dakar confirm a high burden of scabies in their out-patient clinics (Ly, personal communication). We therefore undertook a cluster-randomised, cross-sectional survey employing a standardised diagnostic approach to diagnose scabies and impetigo in peri-urban Dakar.

## 2. Methods

The survey took place in Pikine, a neighbourhood of about 2 km x 3 km in size that lies 10 km outside the centre of Dakar, the capital of Senegal. Pikine is a very densely populated area of single and multi-story concrete and cement brick houses, arranged into a square grid by sand and tarmac streets. The mostly Wolof-speaking, multi-ethnic population is a mix of long-term residents and more recent arrivals. It is considered one of the poorer parts of Dakar but public services such as sanitation, waste removal and piped water are available to most inhabitants.

The border of Pikine was mapped using Google maps. Each block of houses was numbered (N = 530). We excluded 24 blocks of houses containing only large commercial or public buildings (17 blocks) or those, who were too large to be enlisted in a single day by the study team (7 blocks). We then randomly selected 27 housing blocks using simple random selection. The cluster size was selected to allow the study team to be able to visit all households and enlist all household members within the cluster in one full working day.

Two final year post-graduate dermatology students and two second-year post-graduate dermatology students from Université Cheikh Anta Diop Dakar were trained during a 2-day practical training course to diagnose scabies according to the IACS criteria (18). Community health volunteers accompanied the team and helped with translation where this was necessary.

During the household visit basic demographic and socio-economic data and a standardised history and examination for scabies and impetigo was conducted on all participants. The examination involved only the arms up to the axilla, exposed parts of the legs and the face. This has been shown to diagnose 90% of all scabies cases compared to a full body exam in a validation trial. (31) Health-related quality of life data was collected from individuals with scabies. All data was collected using ASUS Zenpad tablets and ODK (Open Data Kit) software.

A case of scabies was defined as fulfilling the 2020 IACS’s criteria of either clinical scabies or suspected scabies. (18) Dermatoscopy or other methods of parasite identification were not available in this community trial to confirm the diagnosis in our participants. The severity of the presentation was classified by the number of lesions present as mild (≤10 lesions), moderate (11-49 lesions) or severe (≥ 50 lesions or crusted scabies). (31) Impetigo was diagnosed on the basis of papular, pustular or ulcerative lesions with associated erythema, crusting, bullae or pus. Severity of impetigo was classified as very mild (≤ 5 lesions), mild (6-10 lesions), moderate (11-49 lesions) or severe (≥ 50 lesions) as described before. (31).

All patients found to have scabies or impetigo were provided treatment free of charge according to standard local treatment guidelines. Scabies was treated with weight based oral ivermectin on day 0 and day 7, or topical benzyl benzoate 10% lotion for children with a weight of less than 15 kg. Impetigo was treated with oral antibiotics for 5 days and wound care. Symptomatic household contacts were also treated. Other moderate or severe dermatological conditions were referred for treatment.

Health-related quality of life was assessed using the Dermatology Quality of Life Index (DLQI) for those 16 years of age or older, the Children’s DLQI (CDLQI) for children aged 7 to 15 years and the Family DLQI (FDLQI) for children 6 years or below. The DLQIs have been used previously for scabies, (32) are validated for use on tablets, (33) and validated translations into French are available. (34) Where French was not the mother tongue of the participant, the questions were read out and translated from French into Wolof by the fieldworkers or the community health volunteers.

Sample size calculations: We assumed that with a design-effect of 1.2 and an average cluster size of 100 we needed to enrol 1600 individuals to detect a prevalence of scabies of 10% with 2% precision. Assuming an average household size of 5 we planned to enrol 16 clusters of 20 households. As the average cluster size was less than 100 participants, we increased the number of randomly selected clusters to 27 to meet our target of 1600 participants.

We calculated the prevalence of scabies and impetigo stratified by demographic variables. For children age was grouped in steps of 5-years until the age of twenty. The remaining adult population was summarised in a single age category. The education of the head of the household was categorised as either no schooling, schooling up to partial secondary school education or at least secondary school education. The type of school was categorised as either Koranic school or any other school. Household size was defined as how many people slept in the house the previous night. The svyset STATA command was used to adjust for the cluster randomized sampling design. For proportions robust standard errors were used to calculate confidence intervals adjusted for clustering at the community level. Logistic regression analysis was done using random-effects regression adjusted for clustering as above. Due to the small number of outcomes univariate modelling was chosen for key socio-demographic variables to avoid overfitting of the model. The relationship between HRQOL and scabies was assessed by calculating the FDLQI, CDLQI and DLQI according to age. Median scores and inter-quartile ranges (IQR) were used to quantify the effect on the QOL. The impact of scabies on the quality of life was assessed as no effect on patient’s life (DLQI scores 0–1), small effect on patient’s life (2-5), moderate effect on patient’s life (6-10), very large effect on patient’s life (11-20) and extremely large effect on patient’s life (21-30). (35) Stata SE 17.0 (StataCorp, College Station, TX) was used for data analysis.

The study was approved by the LSHTM MSc Research Ethics Committee (Ref. No. 27220) and the ‘Comité National d’Éthique pour la Recherche en Santé (CNERS)’ ethics committee in Senegal (Protocol SEN22/03). The study was introduced to and approved by key public health officials in Pikine, the area of study and the ‘Department of Neglected Tropical Diseases’ at the Ministry of Health and Social Action of the Republic of Senegal. Written informed consent was obtained for each subject or from their parent or legal guardian.

## 3. Results

Between July 19^th^ and August 5^th^ 2022 1697 participants from 392 households were enrolled (see figure 1 + 2). The study period fell in the national school holidays and at the start of the rainy season. 1207 (71.1%) participants were female. There was a broadly equal distribution of male and female participants in early childhood but a skewed distribution towards female participants was observed in working-age adults. The median age of the participants was 19 years (IQR 8 – 37 years). The majority of the population were Wolof, Fulani and Serer. The educational attainment for the head of households was generally low with 47.8 % not having attended any formal school and 36.9 % had not completed secondary school. (Table 1)

**Table 1:**
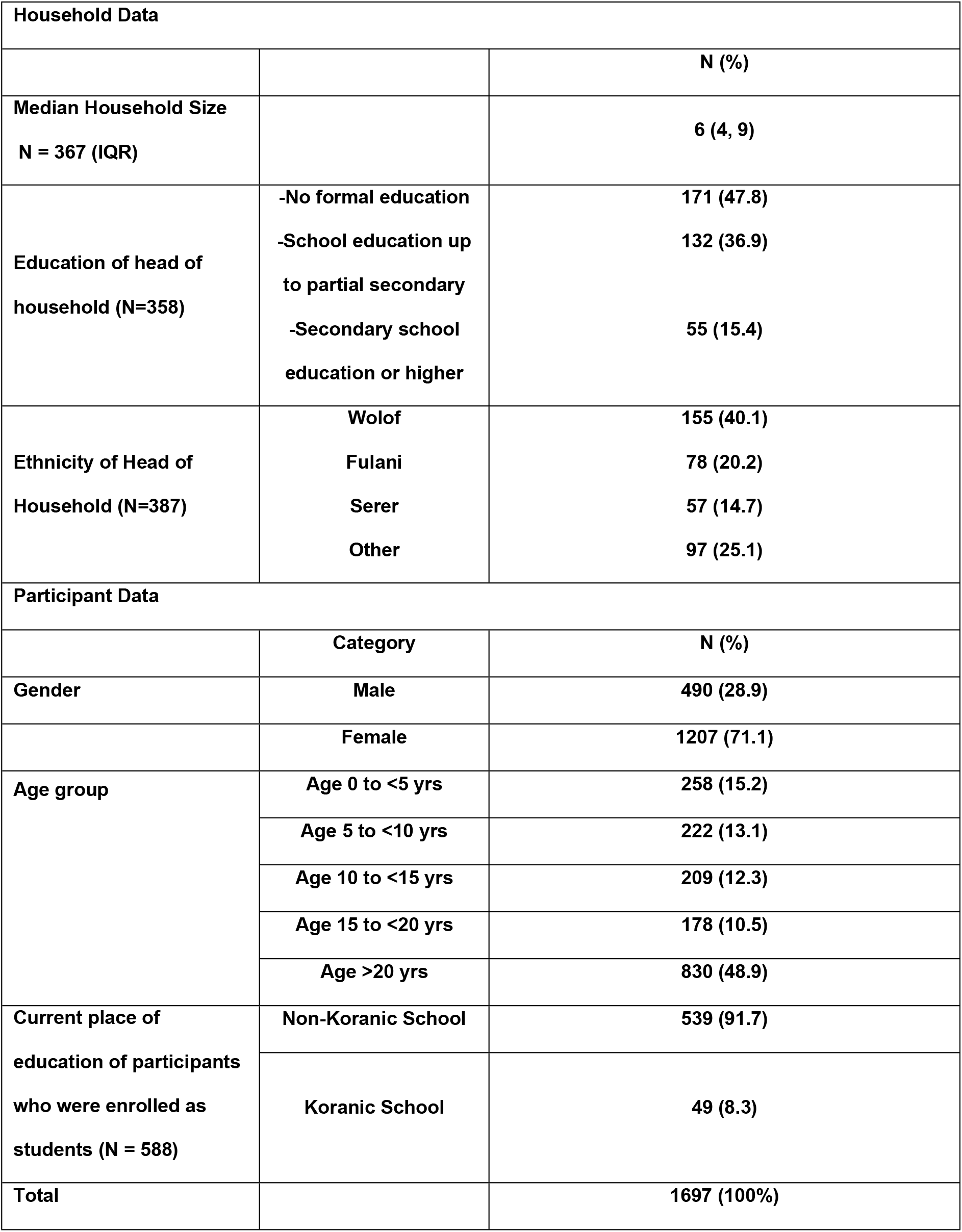
Age and gender of participants

**Fig. 1:**
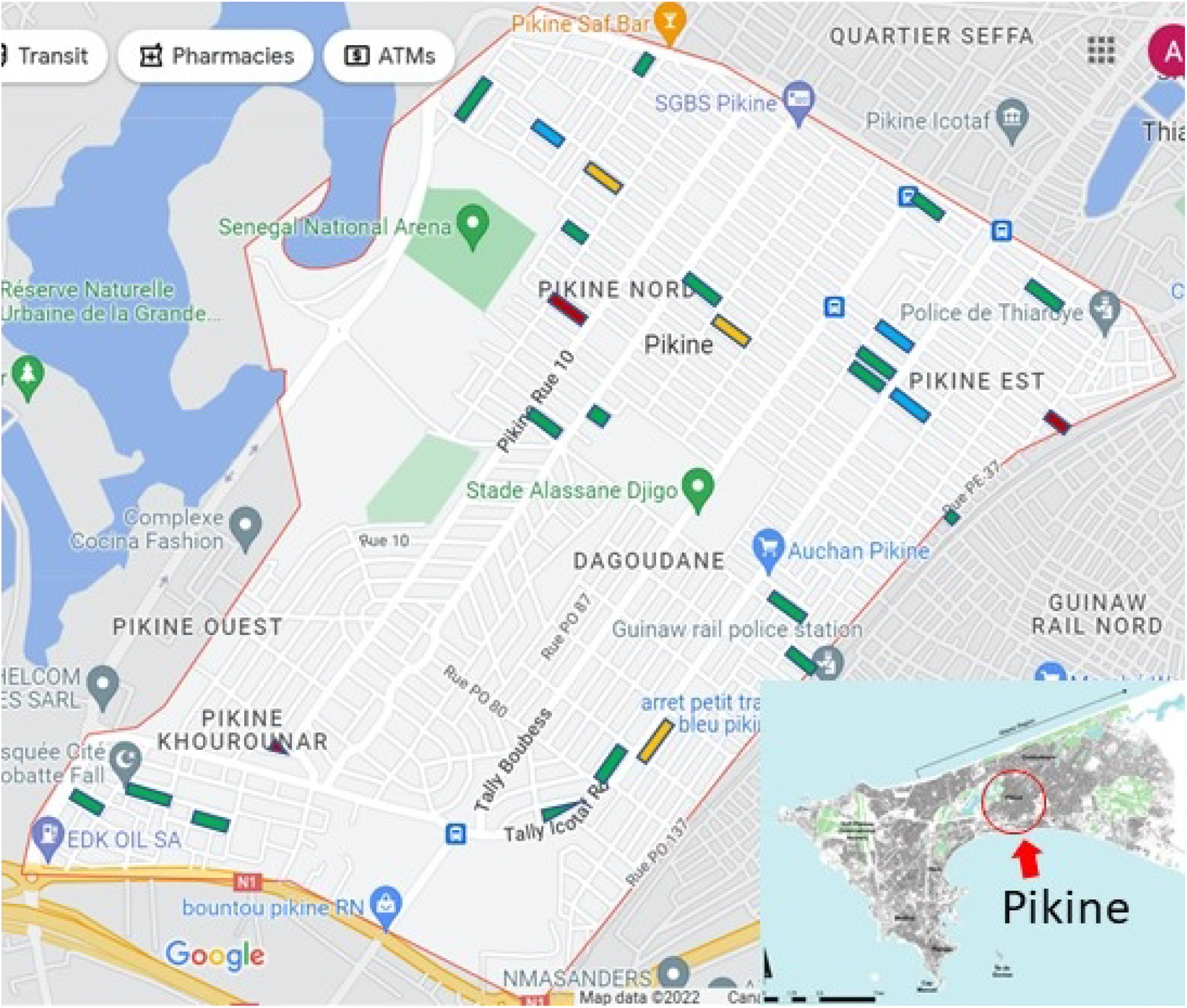
map of Pikine, a suburb of Dakar, the most Western point of Africa, showing the 27 randomly selected clusters, color-coded for different levels of scabies prevalence in the respective clusters, from which participants were recruited: green: no scabies, blue: >0-2% scabies, yellow: >2-4% scabies, red: >4-8.5% scabies

**Fig. 2:**
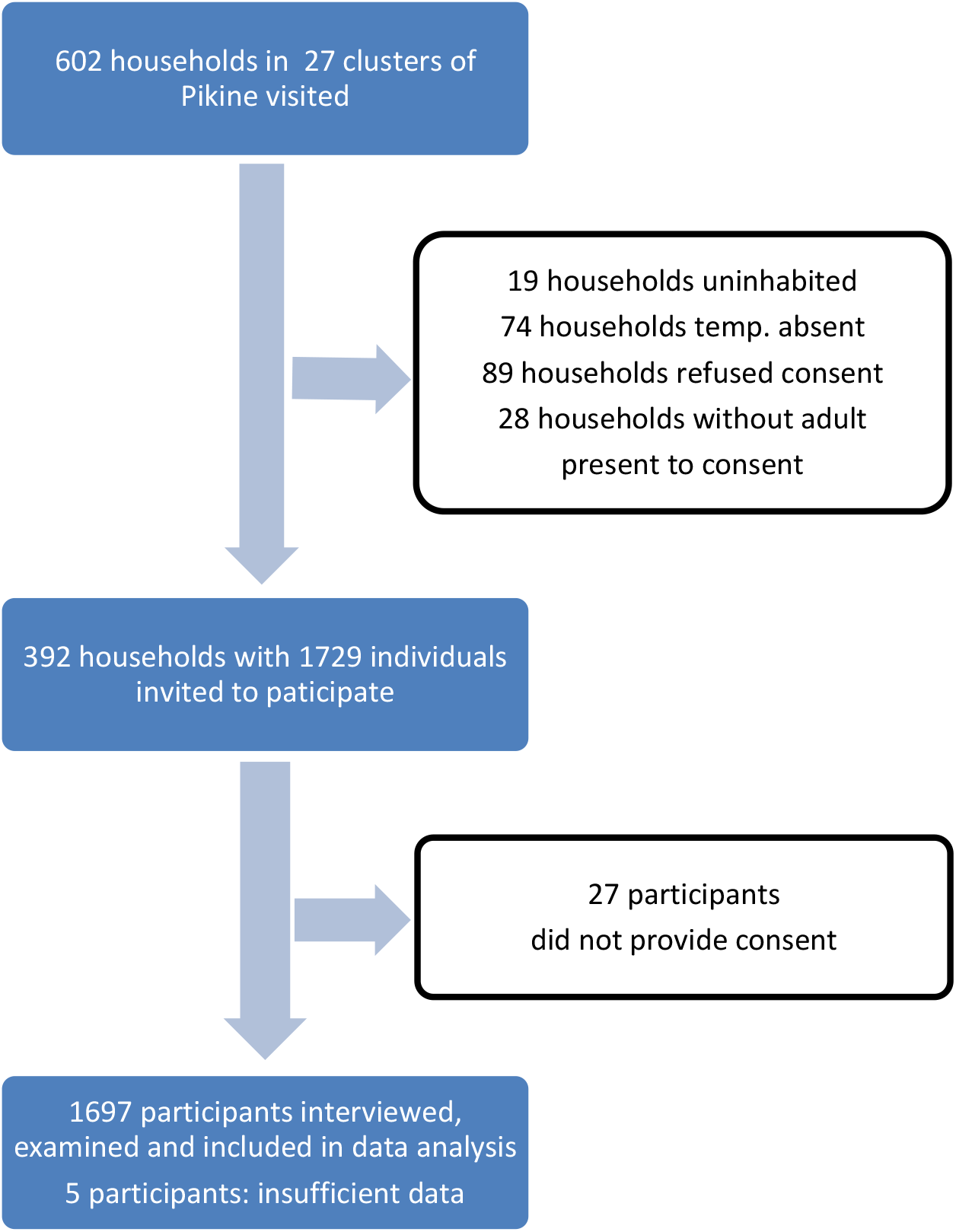
Flow chart of recruitment for scabies and impetigo survey in Pikine, a suburb of Dakar, Senegal

Overall, 27 people (1.6% 95% CI 0.8-3.2%) were diagnosed with scabies. Of these 26 were diagnosed as clinical scabies and one as suspected scabies (C1). Amongst the male population the scabies prevalence was 2.8% compared to 1.1% in females (adjusted odds ratio (OR) 3.1, 95% CI 1.4-6.8, p <0.01). The prevalence of scabies between the 27 clusters ranged from 0% to 8.5%. Amongst the different age categories, scabies was most prevalent amongst school-aged children between the ages of 5 and 10 years (4.5%) compared to 0.4% in the adult population. (Table 3). Of the 27 cases of scabies 11 were mild (40.7%), 3 were of moderate severity (11.1%) and 13 were severe (48.1%).

**Table 3.** Scabies and impetigo prevalence by age category, gender, school type and education of head of household

|  | Participants No (%) | Cases with scabies N (%) | Adjusted odds ratio (95% CI)* | p-value* | Cases with impetigo No (%) | Adjusted odds ratio (95% CI)* | p-value* |
| --- | --- | --- | --- | --- | --- | --- | --- |
| **Age 0-<5 yrs.** | 258 (15.2) | 6 (2.3) | 6.7 (1.6-27.5) | <0.01 | 3 (1.2) | 9.9 (1.0-96.3) | 0.05 |
| **Age 5-<10 yrs.** | 222 (13.1) | 10 (4.5) | 12.1 (3.2-45.4) | <0.01 | 4 (1.8) | 14.2 (1.6-129.7) | 0.02 |
| **Age 10 to <15 yrs.** | 209 (12.3) | 7 (3.3) | 9.1 (2.3-36.4) | <0.01 | 2 (1.0) | 7.8 (0.7-87.8) | 0.10 |
| **Age 15 to <20 yrs.** | 178 (10.4) | 1 (0.6) | 1.5 (0.1-14.4) | 0.74 | 0 (-) | - | - |
| **Age >20 yrs.** | 830 (48.9) | 3 (0.4) | 1 | - | 1 (0) | 1 | - |
| **Total** | **1697** | **27 (1.6)** | **(0.8-3.2)** | **-** | **10 (0.6)** | **(0.3-1.3)** | **-** |
| **Male** | 490 (28.9) | 14 (2.8) | 3.1 (1.4-6.8) | <0.01 | 9 (1.8) | 25 (3.1-200.5) | <0.01 |
| **Female** | 1207 (71.1) | 13 (1.1) | 1 | - | 1 (0%) | 1 | - |
| **Participant attending** | 49 (2.9) | 10 (20.4) | 17.6 (6.0-52.1) | <0.01 | 3 | 16.8 (2.4-118.5) | <0.01 |
| **Koranic school** |  |  |  |  |  |  |  |
| **Participant attending regular school** | 539 (31.8) | 8 (1.5) | 1 | - | 2 | 1 | - |
| **Household head with any education** | 701 (41.3) | 7 (1.0) | 0.5 (0.2-1.3) | 0.16 | 2 (0.3) | 0.61 (0.1-3.3) | 0.57 |
| **Household head without education** | 702 (41.4) | 18 (2.6) | 1 | - | 6 (0.9) | 1 | - |

\* using logistic regression analysis corrected for cluster sampling design

A total of 10 participants were diagnosed with impetigo (0.6%, 95% CI 0.3-1.3%). Of these 5 also had scabies. The severity of impetigo was classified as very mild in 5 cases and mild in the remaining 5 cases. Overall, 9 of the 10 cases (90%) were diagnosed in boys.

The prevalence of scabies was higher amongst children attending koranic schools compared to non-koranic schools (20.4% vs 1.5%, OR 17.6 95% CI 6.0-52.1, p <0.01). There was no significant association between the education of household heads and the presence of scabies or impetigo in participants (Table 3). Household size was not associated with scabies.

For participants who completed a DLQI the median score was 10.00 (IQR 8-12), for participants who completed a CDLQI the median score was 8.75 (IQR 4.5-13) and for participants where a FDLQI score was completed the median was 5.83 (IQR 2-10). As the number of cases was small we did not stratify by disease severity.

## 4. Discussion

In this study we found a relatively low prevalence of scabies and impetigo in a peri-urban region of Dakar. Cases of scabies were not evenly distributed in the population and were much more common in school-aged male children, in particular those who attended Koranic schools. These data suggest that transmission of scabies in the general population is relatively limited and that most cases occur from transmission within specific facilities or locations such as schools, e.g. Koranic.

The prevalence of scabies seen in this study are lower than that reported in a survey in Monrovia in Liberia and a second survey in Sukuta in The Gambia. (20)(29) Both of these surveys had a prevalence of scabies that approached 10% in the general population suggesting that in these settings there was more generalized transmission of scabies. In the absence of any recent health interventions to reduce the burden of disease in Pikine, there are several possible reasons for this difference. First, the GDP per capita in Senegal is about twice that of The Gambia and three times as high compared to Liberia. Second, scabies is associated with crowding and it is conceivable that living conditions in Pikine are less crowded. Third, it may be that the population in Pikine has better access to preventive or therapeutic health services than inhabitants in Monrovia or Sukuta. In addition, there may be other unknown geographic, seasonal, socio-economic or public health reasons for the low burden of disease which we were unable to explore.

We found that the health-related QOL impact of scabies on peoples’ lives was mild to moderate which is broadly in line with previous studies. (36)(8)(37) As the total number of cases of scabies was low, our ability to assess relationships between disease severity and quality of life measures was limited.

Compared to the overall study population we found a very high rate of scabies infection in pupils attending Koranic schools compared to attendants of non-Koranic schools. As a result, individuals attending Koranic schools made up more than one third of all cases with scabies despite representing less than 3% of participants. We have previously shown a high prevalence of scabies amongst children at Koranic schools. (30) The increased rate of scabies seen in male participants likely reflects this increased burden amongst attendees at Koranic schools. Our data suggest targeted interventions may be the most effective strategy to reduce the burden of scabies in these settings.

Our study has a number of strengths including a large sample size, the use of a robust cluster-randomised sampling approach and the use of standardized examinations using the IACS criteria performed by well trained staff. The entire period of recruitment fell into the school summer holidays in Senegal, which allowed us to recruit a larger part of the school-aged children at home. There are also a number of limitations, given the nature of the fieldwork, we did not perform dermatoscopy or other direct visualisation methods. However, the performance of the clinical IACS criteria has previously been shown to be acceptable at the population level and we do not think this will have impacted our findings. We undertook a cross-sectional survey and cannot exclude that seasonal variations might occur as has been shown for impetigo in The Gambia (29). Despite best efforts to enroll all members of each household recruitment took place during working hours only, resulting in an under representation of adult men, which has also been seen in other studies. (20)

In summary we found a relatively low overall prevalence of scabies in the general population but evidence of much higher levels of transmission in particular environments, notably Koranic schools. Our data highlight the need for scabies prevalence surveys to consider not only the overall population prevalence but also specific sub-populations where targeted interventions may be required. Further studies specifically involving Koranic schools, including intervention studies should be considered.

## Data Availability

A data set has been uploaded and attached to this application under supporting documents.

## Acknowledgements

- We thank all families who participated in the study.
- Mrs. Awa Dieye, Mrs. Nogaye Diouf, Mrs. Marema Ndiaye, Mr. Mame Thiano Seye and Mrs. Aissatou Gaye Fall, community health workers and matron at Centre de Santé Dominique in Pikine, helped with the conduct of the field work and translations.
- Chrissy Roberts from the London School of Hygiene and Tropical Medicine Global Health Analytics kindly provided electronic data solutions.

## Reference List

1. Vos T, Allen C, Arora M, Barber RM, Brown A, Carter A, et al. Global, regional, and national incidence, prevalence, and years lived with disability for 310 diseases and injuries, 1990–2015: a systematic analysis for the Global Burden of Disease Study 2015. Lancet. 2016;388(10053):1545–602.

2. Vos T, Abajobir AA, Abbafati C, Abbas KM, Abate KH, Abd-Allah F, et al. Global, regional, and national incidence, prevalence, and years lived with disability for 328 diseases and injuries for 195 countries, 1990-2016: A systematic analysis for the Global Burden of Disease Study 2016. Lancet. 2017;390(10100):1211–59.

3. WHO. Ending the neglect to attain the Sustainable Development Goals: a road map for neglected tropical diseases 2021–2030. Word Health Organization. 2020. 196 p.

4. Romani L, Steer AC, Whitfeld MJ, Kaldor JM. Prevalence of scabies and impetigo worldwide: A systematic review. Lancet Infect Dis [Internet]. 2015;15(8):960–7. Available from: http://dx.doi.org/10.1016/S1473-3099(15)00132-2

5. Gibbs S. Skin disease and socioeconomic conditions in rural Africa: Tanzania. Int J Dermatol. 1996;35(9):633–9.

6. Chosidow O. Scabies. N Engl J Med. 2006;6(354):1718–27.

7. Heukelbach J, Feldmeier H. Scabies. Lancet. 2006;367(9524):1767–74.

8. Worth C, Heukelbach J, Fengler G, Walter B, Liesenfeld O, Feldmeier H. Impaired quality of life in adults and children with scabies from an impoverished community in Brazil. Int J Dermatol. 2012;51(3):275–82.

9. Thean LJ, Jenney A, Engelman D, Romani L, Wand H, Mudaliar J, et al. Hospital admissions for skin and soft tissue infections in a population with endemic scabies: A prospective study in Fiji, 2018-2019. PLoS Negl Trop Dis [Internet]. 2020;14(12):e0008887. Available from: http://dx.doi.org/10.1371/journal.pntd.0008887

10. Whittle HC, Abdullahi MT, Fakunle F, Parry EHO, Rajkovic AD, Ibrahim A, et al. Scabies, pyoderma and nephritis in Zaria, Nigeria. A clinical and epidemiological study. Trans R Soc Trop Med Hyg. 1973;67(3):349–63.

11. Svartman M, Finklea JF, Earle DP, Potter EV P-KT. Epidemic scabies and acute glomerulonephritis in Trinidad. Lancet. 1972;1:249–51.

12. Marshall CS, Cheng AC, Markey PG, Towers RJ, Richardson LJ, Fagan PK, et al. Acute post-streptococcal glomerulonephritis in the Northern Territory of Australia: A review of 16 years data and comparison with the literature. Am J Trop Med Hyg. 2011;85(4):703–10.

13. Thornley S, Marshall R, Jarrett P, Sundborn G, Reynolds E, Schofield G. Scabies is strongly associated with acute rheumatic fever in a cohort study of Auckland children. J Paediatr Child Health. 2018;54(6):625–32.

14. Thornley S, King R, Marshall R, Oakley A, Sundborn G, Harrower J, et al. How strong is the relationship between scabies and acute rheumatic fever? An analysis of neighbourhood factors. J Paediatr Child Health. 2020;56(4):600–6.

15. WHO. Report of the Tenth Meeting of the WHO Strategic and Technical Advisory Group for Neglected Tropical Diseases [Internet]. Geneva; 2017. Available from: https://www.who.int/docs/default-source/ntds/strategic-and-advisory-group-on-neglected-tropical-diseases-(stag-ntds)/tenth-ntd-stag-report-2017.pdf?sfvrsn=9ec99065_2

16. Engelman D, Cantey PT, Marks M, Solomon AW, Chang AY, Chosidow O, et al. The public health control of scabies: priorities for research and action. Lancet [Internet]. 2019;394(10192):81–92. Available from: http://dx.doi.org/10.1016/S0140-6736(19)31136-5

17. Engelman D, Fuller LC, Steer AC. Consensus criteria for the diagnosis of scabies: A Delphi study of international experts. PLoS Negl Trop Dis [Internet]. 2018;12(5):1–9. Available from: http://dx.doi.org/10.1371/journal.pntd.0006549

18. Engelman D, Yoshizumi J, Hay RJ, Osti M, Micali G, Norton S, et al. The 2020 International Alliance for the Control of Scabies Consensus Criteria for the Diagnosis of Scabies. Br J Dermatol. 2020;183(5):808–20.

19. Amoako YA, Phillips RO, Arthur J, Abugri MA, Akowuah E, Amoako KO, et al. A scabies outbreak in the north east region of Ghana: The necessity for prompt intervention. PLoS Negl Trop Dis [Internet]. 2020;14(12):1–12. Available from: http://dx.doi.org/10.1371/journal.pntd.0008902

20. Collinson S, Timothy J, Zayzay SK, Kollie KK, Lebas E, Candy N, et al. The prevalence of scabies in Monrovia, Liberia: A population-based survey. PloS Negl Trop Dis [Internet]. 2020;14(12):e0008943. Available from: http://dx.doi.org/10.1371/journal.pntd.0008943

21. Walker SL, Collinson S, Timothy J, Zayay S, Kollie KK, Lebas E, et al. A community-based validation of the international alliance for the control of scabies consensus criteria by expert and non-expert examiners in liberia. PLoS Negl Trop Dis [Internet]. 2020;14(10):1–11. Available from: http://dx.doi.org/10.1371/journal.pntd.0008717

22. Romani L, Koroivueta J, Steer AC, Kama M, Kaldor JM, Wand H, et al. Scabies and Impetigo Prevalence and Risk Factors in Fiji: A National Survey. PLoS Negl Trop Dis. 2015;9(3):1–10.

23. Mason DS, Marks M, Sokana O, Solomon AW, Mabey DC, Romani L, et al. The Prevalence of Scabies and Impetigo in the Solomon Islands: A Population-Based Survey. PLoS Negl Trop Dis. 2016;10(6):1–10.

24. M Harris 1, D Nako, T Hopkins, D M Powell, C Kenny, C Carroll KC. Skin infections in Tanna, Vanuatu in 1989. P N G Med J. 1992;35(2):137–43.

25. R J Eason TT-J. Resurgent yaws and other skin diseases in the Western Province of the Solomon Islands. P N G Med J. 1985;28(4):247–50.

26. Galván-Casas C, Mitjá O, Esteban S, Kafulafula J, Phiri T, Navarro-Fernández í, et al. A facility and community-based assessment of scabies in rural malawi. PLoS Negl Trop Dis. 2021;15(6):1–11.

27. Misganaw B, Nigatu SG, Gebrie GN, Kibret AA. Prevalence and determinants of scabies among school-age children in Central Armachiho district, Northwest, Ethiopia. PLoS One [Internet]. 2022;17(6):e0269918. Available from: http://dx.doi.org/10.1371/journal.pone.0269918

28. Marks M, Sammut T, Cabral MG, Silva ET Da, Goncalves A, Rodrigues A, et al. The prevalence of scabies, pyoderma and other communicable dermatoses in the Bijagos Archipelago, Guinea-Bissau. PLoS Negl Trop Dis. 2019;13(11):1–9.

29. Armitage EP, Senghore E, Darboe S, Barry M, Camara J, Bah S, et al. High burden and seasonal variation of paediatric scabies and pyoderma prevalence in the Gambia: A cross-sectional study. PLoS Negl Trop Dis [Internet]. 2019;13(10):1–16. Available from: http://dx.doi.org/10.1371/journal.pntd.0007801

30. Ly F, Faye A, Wone I, Lelo S, Diouf A, Koundio A, et al. Scabies in Koranic schools in Dakar, Senegal: Prevalence and risk factors. Int J Dermatology, Venereol Lepr Sci. 2021;4(2):49–54.

31. Marks M, Engelman D, Romani L, Mason D, Sokana O, Kama M, et al. Exploration of a simplified clinical examination for scabies to support public health decision-making. PLoS Negl Trop Dis [Internet]. 2018;12(12):1–9. Available from: http://dx.doi.org/10.1371/journal.pntd.0006996

32. Thanushah B, Nilaweera A, Junejo MH, Patel J, Oyebanji O. GD07: The use of the Dermatology Life Quality Index suite of patient-reported outcome measures in neglected tropical diseases of the skin: a systematic review. Br J Dermatol. 2022;187(S1):179–80.

33. Ali FM, Johns N, Finlay AY, Salek MS, Piguet V. Comparison of the paper-based and electronic versions of the Dermatology Life Quality Index: evidence of equivalence. Br J Dermatol. 2017;177(5):1306–15.

34. No Title [Internet]. Available from: https://www.cardiff.ac.uk/medicine/resources/quality-of-life-questionnaires/dermatology-life-quality-index#:~:text = The DLQI is calculated by,affected by their skin disease.

35. Hongbo Y, Thomas CL, Harrison MA, Salek MS, Finlay AY. Translating the science of quality of life into practice: What do dermatology life quality index scores mean? J Invest Dermatol [Internet]. 2005;125(4):659–64. Available from: http://dx.doi.org/10.1111/j.0022-202X.2005.23621.x

36. Walker SL, Lebas E, De Sario V, Deyasso Z, Doni SN, Marks M, et al. The prevalence and association with health-related quality of life of tungiasis and scabies in schoolchildren in southern Ethiopia. PLoS Negl Trop Dis. 2017;11(8):1–11.

37. Lake SJ, Engelman D, Sokana O, Nasi T, Boara D, Marks M, et al. Healthrelated quality of life impact of scabies in the Solomon Islands. Trans R Soc Trop Med Hyg. 2022;116(2):148–56.

